# Parental infertility and offspring cardiometabolic trajectories up to 25 years: a pooled analysis of three European cohorts

**DOI:** 10.1101/2023.10.10.23296797

**Authors:** Álvaro Hernáez, Ahmed Elhakeem, Henrique Barros, Tanja G. M. Vrijkotte, Abigail Fraser, Deborah A. Lawlor, Maria C. Magnus

## Abstract

**Objective:** To assess whether parental infertility is associated with differences in cardiometabolic trajectories in offspring from childhood to 25 years of age.

**Design:** Pooled analysis of three European pregnancy cohort studies.

**Subjects:** Up to 14,609 singletons from three pregnancy cohorts (the UK Avon Longitudinal Study of Parents and Children, the Portuguese Geraçao 21, and the Amsterdam Born Children and their Development study).

**Exposure:** Parental infertility defined as time-to-pregnancy ≥12 months.

**Main Outcome Measures:** Trajectories of body mass index (BMI), waist circumference, systolic blood pressure (SBP), diastolic blood pressure (DBP), low-density lipoprotein cholesterol (LDL-C), high-density lipoprotein cholesterol (HDL-C), triglycerides, and glucose from childhood to 25 years of age were compared in offspring of couples with and without infertility. Trajectories were modelled using mixed-effects models with natural cubic splines adjusting for cohort, sex of the offspring, and maternal factors (age, body mass index, smoking, educational level, parity, and ethnicity). Predicted levels of cardiometabolic traits up to 25 years of age were compared by parental infertility.

**Results:** Offspring of couples with infertility had increasingly higher BMI (difference in mean predicted levels by age 25: +1.09 kg/m^2^, 95% confidence interval [0.68 to 1.50]) and suggestively higher DBP at age 25 (+1.21 mmHg [0.00 to 2.43]). Their LDL-C tended to be higher, and their HDL-C values tended to be lower over time (age 25, LDL-C: +4.07% [-0.79 to 8.93]; HDL-C: −2.78% [-6.99 to 1.43]). At middle-late adolescence, offspring of couples with infertility had higher waist circumference (age 17: +1.05 cm [0.11 to 1.99]) and SBP (age 17: +0.93 mmHg [0.044 to 1.81]), but these differences attenuated at later ages. No clear inter-group differences in triglyceride and glucose trajectories were observed. Further adjustment for paternal age, body mass index, smoking, and educational level, and both parent’s history of diabetes and hypertension in the cohort with this information available (Avon Longitudinal Study of Parents and Children) did not attenuate inter-group differences.

**Conclusion:** Offspring of couples with infertility have increasingly higher BMI over the years, suggestively higher blood pressure levels, and tend to have greater values of LDL-C and lower values of HDL-C with age.

## INTRODUCTION

Cardiovascular disease is more incident among individuals reporting fertility problems (1–6). Women with infertility have higher body mass index (BMI), waist circumference (WC), and triglycerides, greater odds of diabetes, and lower concentrations of high-density lipoprotein cholesterol (HDL-C) compared to fertile women (7), and men with infertility are more prone to be obese and hypertensive (8). A more adverse cardiometabolic health could also be present in their offspring. First, cardiometabolic disease and its risk factors are inheritable (9).

Second, infertility is linked to pregnancy complications such as gestational diabetes (10) and hypertensive disorders of pregnancy (11), which are associated with increased cardiometabolic risk in the offspring (12, 13). Third, couples with fertility problems are likely to use assisted reproductive technologies (ART), which are related to differences in cardiometabolic trajectories in the offspring (14, 15). We previously reported that ART-conceived offspring had a lower BMI, WC, and systolic and diastolic blood pressure (SBP and DBP, respectively) in childhood, and subtle trajectories to higher SBP and triglycerides in young adulthood, although these differences were small (14, 15). Only three other studies to date have examined cardiometabolic risk factors in offspring of individuals with infertility (16–18), most focused on changes in BMI, and reported minimal or null differences between offspring of couples with and without infertility. Whether differences in cardiometabolic trajectories of offspring conceived by ART are due to ART procedures or underlying parental infertility remains unclear.

Our study therefore aims to determine the association of parental infertility (not considering ART use) with offspring trajectories of BMI, WC, SBP, DBP, LDL-C, HDL-C, triglycerides, and glucose from birth to adulthood.

## METHODS

### Population description

Our analyses are part of the Assisted Reproductive Technology and Health (A.R.T-HEALTH) partnership (14, 15). All cohorts in the partnership were invited to participate in this study, with the inclusion criteria being that they had information on time to pregnancy and two or more measurements taken at different times for at least one of the eight cardiometabolic traits of interest. Three cohorts fulfilled these requirements: the UK Avon Longitudinal Study of Parents and Children [ALSPAC; birth years: 1990-1992] (19–23), the Portuguese Geraçao 21 [G21; birth years: 2000-2006] (24), and the Dutch Amsterdam Born Children and their Development Study [ABCD; birth years 2003-2004] (25). A detailed description of the number of mothers, partners, pregnancies, and offspring included ALSPAC is available in the **Supplementary Materials**. ALSPAC study data were collected and managed using Research Electronic Data Capture electronic data capture tools hosted at the University of Bristol (26). The ALSPAC study website (http://www.bristol.ac.uk/alspac/researchers/our-data/) contains details of all the data that is available through a searchable data dictionary and variable search tool.

Figure 1 shows the flow chart of the study. Our analyses considered singletons only and are reported following the guidelines described by the STrengthening the Reporting of OBservational studies in Epidemiology statement.

**Figure 1.**
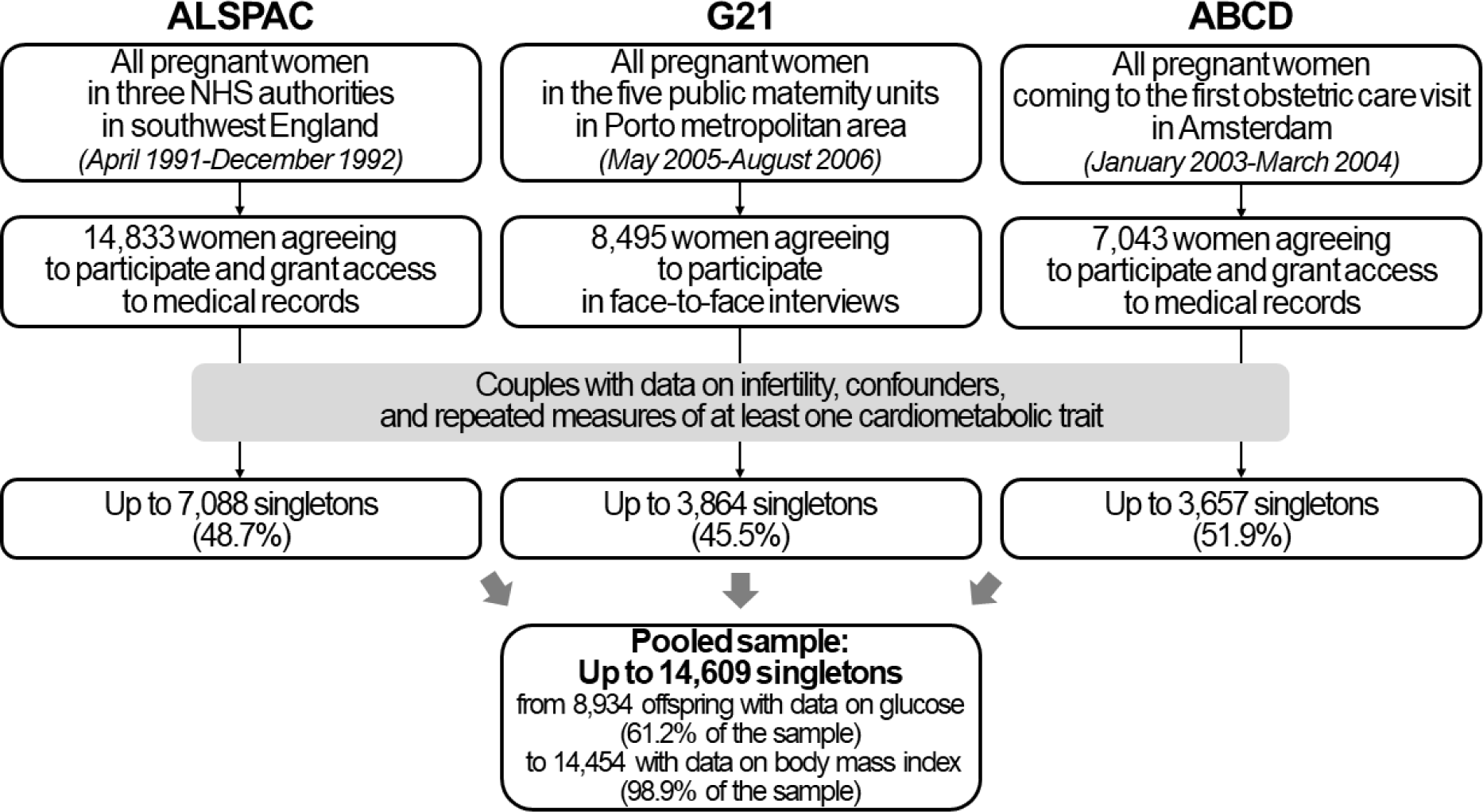
Flow chart of the study

### Infertility

The three cohorts asked the mothers how long (in months) they tried to conceive before becoming pregnant. Infertility was defined as a time-to-pregnancy greater than 12 months. Offspring of couples reporting a time-to-pregnancy ≤12 months (“fertile”) were included in the reference group. Unplanned pregnancies were included in the reference group. ART-conceived offspring were excluded from the analyses.

### Cardiometabolic risk factors in the offspring

ALSPAC collected data of BMI (median number of measures: 10 [interquartile range-IQR-9]; age range: 0 to 26 years), WC (median: 7 measures [IQR 4]; 2-26 years), blood pressure (median: 7 measures [IQR 4]; 3-26 years), lipid profile (median: 2 measures [IQR 3]; 3-26 years) and glucose (median: 2 measures [IQR 2]; 7-26 years). G21 collected data of BMI (median: 18 measures [IQR 9]; 0-12 years) and the rest of risk factors (up to 3 measures; 3-12 years). ABCD collected data of BMI (median: 11 measures [IQR 4]; 0-12 years) and the rest of risk factors (up to 2 measures; 5-13 years). The combined dataset had repeated measures of BMI (median: 12 measures [IQR 7]), WC (median: 3 measures [IQR 4]), blood pressure (median: 3 measures [IQR 5]), lipid profile (median: 2 measures [IQR 2]), and glucose (median: 2 measures [IQR 1]). The number of participants with cardiometabolic trait data at each visit is available in **Supplemental Table 1**.

In ALSPAC, height and weight come from parental questionnaires (before age 16), self-measurements (after age 16), health records, school records, or research clinics, while waist circumference was measured to the nearest mm at research clinics. In G21, anthropometric data was obtained by trained examiners. In ABCD, anthropometric information came from the Youth Health Care registration of the Public Health Service in Amsterdam (by trained nurses) or research clinics. Blood pressure was measured at rest with validated sphygmomanometers (ALSPAC: average of 2 measurements; G21: average of 2 measurements if the difference between them was <5 mmHg [if greater, a third measurement was conducted and the mean of the two closest ones was calculated]; ABCD: average of 2 measurements). Regarding the biochemical determinations, in ALSPAC, lipids were measured by standard laboratory tests (following the standard Lipid Research Clinics Protocol, using enzymatic reagents) in non-fasting plasma/serum up to age 9 and in fasting samples for older ages, and glucose was quantified in non-fasting venous blood samples by nuclear magnetic resonance at age 7 and by standard laboratory tests in fasting samples at older ages (as above). In G21, lipids and glucose were assessed in fasting venous blood samples (lipids: enzymatic colorimetric assays; glucose: hexokinase method). In ABCD, lipids and glucose were measured using a validated ambulatory collection kit on fasting capillary blood samples (Demecal: Lab Anywhere, Haarlem, the Netherlands). LDL-C was calculated using the Friedewald formula (LDL-C = total cholesterol – HDL-C – [triglycerides/5]).

### Confounders

Prior to analyses, we defined confounders as known/plausible causes of parental infertility and offspring cardiometabolic traits: parental age, socioeconomic position (education as a proxy), ethnicity, parity/number of children, and pre-pregnancy/early-pregnancy cardiometabolic risk factors of the parents (BMI, smoking, hypertension, diabetes) (14, 15). As some were assessed during pregnancy, we assumed a strong correlation between pre-pregnancy and during-pregnancy measures. As we pooled individual participant data into one database, we only included confounders that were available in all studies and harmonized them to the lowest common denominator (14, 15). These were: maternal age at pregnancy/birth (years, continuous), pre-pregnancy or early pregnancy maternal BMI (kg/m^2^, continuous), pre-pregnancy or early pregnancy maternal smoking (yes/no), maternal educational level (medium-high level/low level; medium-high education level was defined as 13+ years [ALSPAC], 10+ years [G21], and 14+ years of schooling [ABCD], while low education level was defined as any shorter duration of schooling), parity (0/1 or more), and ethnicity/country of birth (European/other). We also adjusted for offspring sex to improve estimate precision. ALSPAC had further information on paternal pre-pregnancy age (continuous), early-pregnancy BMI (continuous), history of smoking (yes/no), and educational level (medium-high level/low level, as described above), and both parents’ history of diabetes and hypertension (yes/no).

### Ethics

All cohorts comply with the Declaration of Helsinki for Medical Research involving Human Subjects. In all three studies informed written consent for the collection and use of data was provided by parents up to offspring age 16, 12, and 13 (in ALSPAC, G21, and ABCD, respectively) and thereafter by the offspring in ALSPAC. in ALSPAC, ethical approval is provided by the ALSPAC Ethics and Law Committee and UK Local Research Ethics Committees (http://www.bristol.ac.uk/alspac/researchers/research-ethics/), consent for biological samples was collected following the Human Tissue Act (2004), and informed consent for the use of data collected via questionnaires and clinics was obtained following the recommendations of the ALSPAC Ethics and Law Committee at the time. In G21, ethical approval is provided by the Ethics Committee of Hospital de São João and the Institute of Public Health of the University of Porto. In ABCD, it is provided by the Netherlands Central Committee on Research involving Human Subjects, the Medical Ethical Committees of the participating hospitals, and the Registration Committee of the Municipality of Amsterdam.

### Statistical analyses

We described normally distributed variables with means and standard deviations and categorical variables with proportions. Differences in baseline characteristics of participants included and not included in our analyses were studied using t-tests and chi-squared tests for normally distributed continuous variables and categorical variables, respectively.

Individual participant data was merged and analyzed together. We assessed the association of parental infertility with offspring age-related cardiometabolic trajectories using mixed effects linear regression with natural cubic splines (27). We used a natural cubic spline function for age as a fixed effect to model the cardiometabolic health trajectories, which allowed for the detection of non-linear changes in outcomes with age. Models included seven knots for BMI, five for WC, three for SBP and DBP, two for lipid profile measures, and one for glucose (14, 15). Knots were placed at equal quantiles of age. Each cohort contributed data for different age ranges (ALSPAC up to age 26, G21 up to 12, and ABCD up to 13). Main analyses were adjusted for cohort, offspring sex, maternal age, maternal BMI, maternal smoking, maternal educational level, parity, and ethnicity, and included an interaction term between age at cardiometabolic measurements and parental infertility to allow for different trajectories in offspring born of couples with infertility and those without. We used predicted values to plot mean trajectories in both populations and used them to calculate the mean differences in cardiometabolic factors between ages 1 and 25 every 2 years. Differences in the measured values are presented for BMI, WC, SBP and DBP. LDL-C, HDL-C, triglycerides, and glucose were log-transformed prior to analysis and inter-group differences presented are the percent difference between the exposure groups. Participants with missing covariates were excluded from the analyses.

As we were unable to adjust for all a priori defined confounders in the main analyses, in the subsample of ALSPAC participants with available data on the confounders in the main analyses plus the additional confounders (paternal age, BMI, smoking, and educational level, and both parent’s history of diabetes and hypertension, *n* = 3,847), we compared the analyses adjusted for the covariates available in all three cohorts and those adjusted for all the confounders. Besides, we conducted three sensitivity analyses. First, to check the consistency of trajectories across cohorts, we conducted trajectory analyses in ALSPAC, G21 and ABCD separately and compared their shapes by visual inspection. Second, to check the assumption that the results are from the same underlying population, we limited our analyses to the age range for which there is information in the three cohorts (ages 1-15 for BMI, ages 5-11 for the other outcomes) and compared their shapes with the main results by visual inspection. Finally, to check whether our results were consistent with a broader definition of infertility (time-to-pregnancy >12 months + ART use), we repeated the analyses using the definition for the exposure that also included offspring conceived by ART.

All analyses were performed in R Software version 4.0.3.

## RESULTS

### Study population

We studied up to 14,609 singleton offspring of whom 1,351 (9.5%) were born to couples with infertility. Maternal age at birth, BMI and smoking were similar across cohorts. Nevertheless, low maternal education and high parity were more frequent in ALSPAC, and a higher proportion of participants of non-European origin was observed in ABCD (**Table 1**).

**Table 1.**
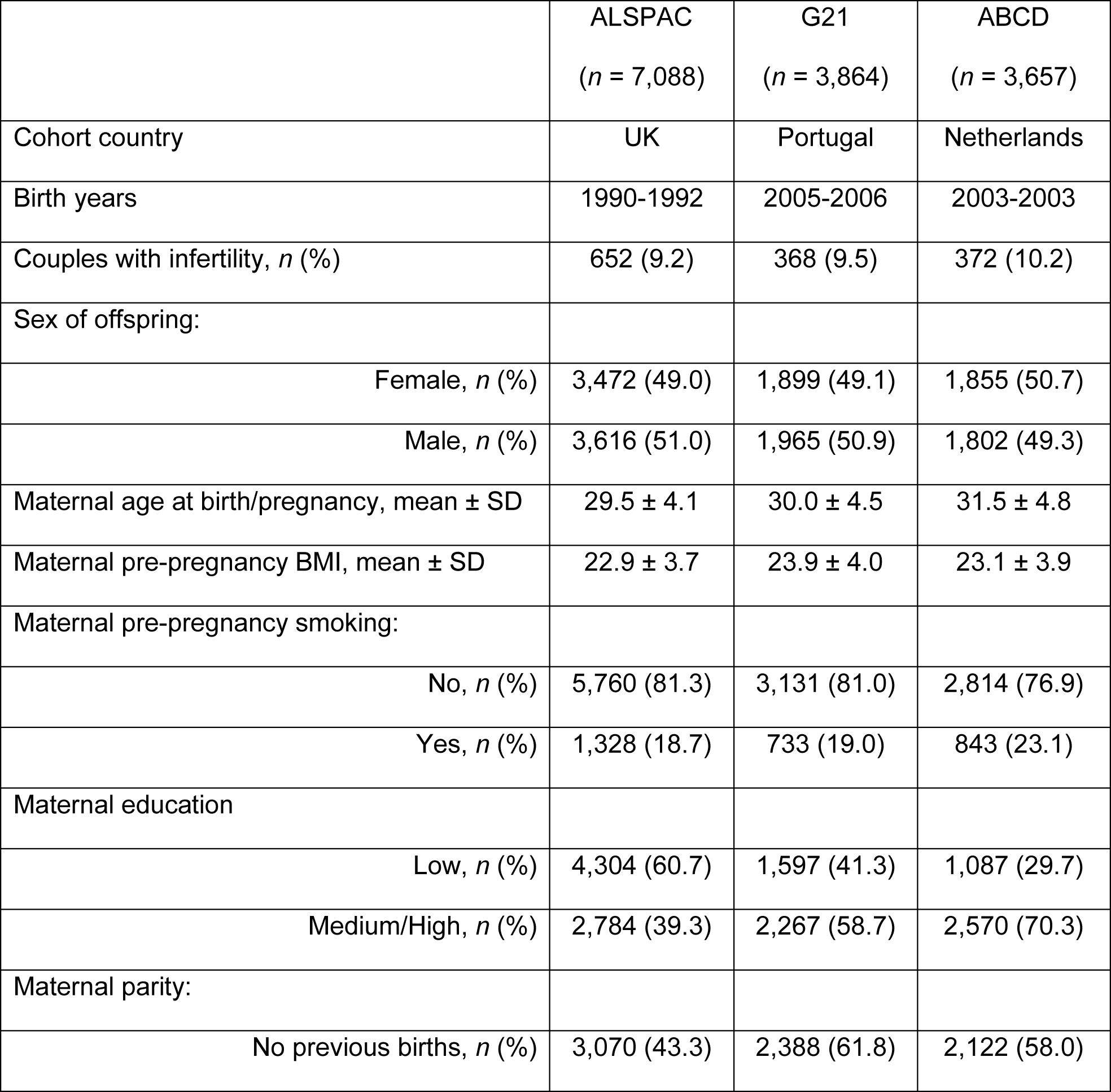
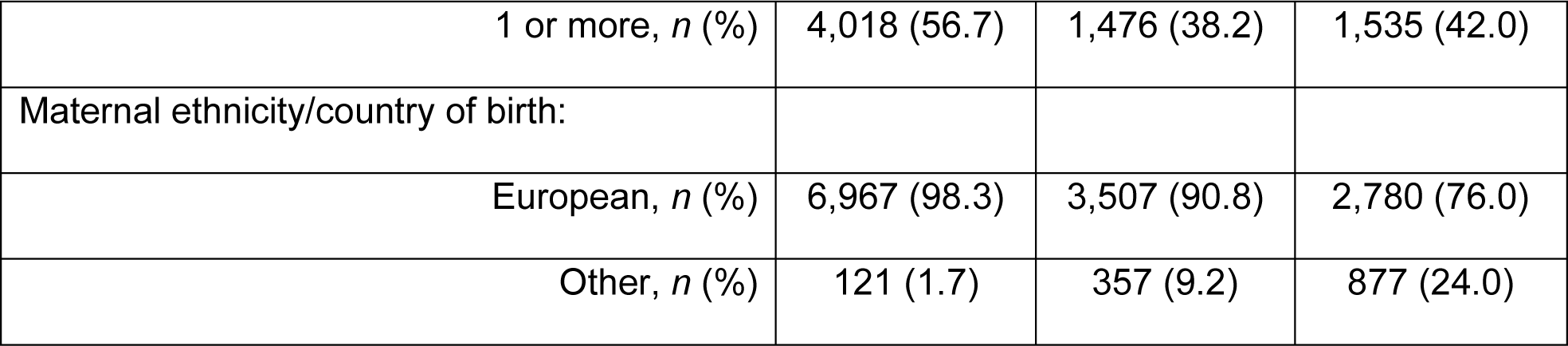
Participant characteristics by cohort.

Parents of non-included offspring were younger, more prone to be smokers (in ALSPAC and ABCD), less educated (in ALSPAC and G21), more likely to be of non-European origin, and more likely to have more than one offspring (in G21 and ABCD) (**Supplemental Table 2**).

### Adiposity trajectories

In the adjusted analyses with pooled individual participant data, BMI differences emerged in childhood and increased with age (Figure 2A). At age 25, offspring of couples with infertility had higher mean BMI (+1.09 kg/m^2^, 95% CI 0.68 to 1.50, Figure 2B). Offspring of couples with infertility presented lower mean WC during childhood (age 5: −0.39 cm, 95% CI −0.90 to 0.12), which evolved to higher values during adolescence (age 17: +1.05 cm, 95% 0.11 to 1.99), and these differences were attenuated in adulthood (**Figures 2C** and **2D**).

**Figure 2.**
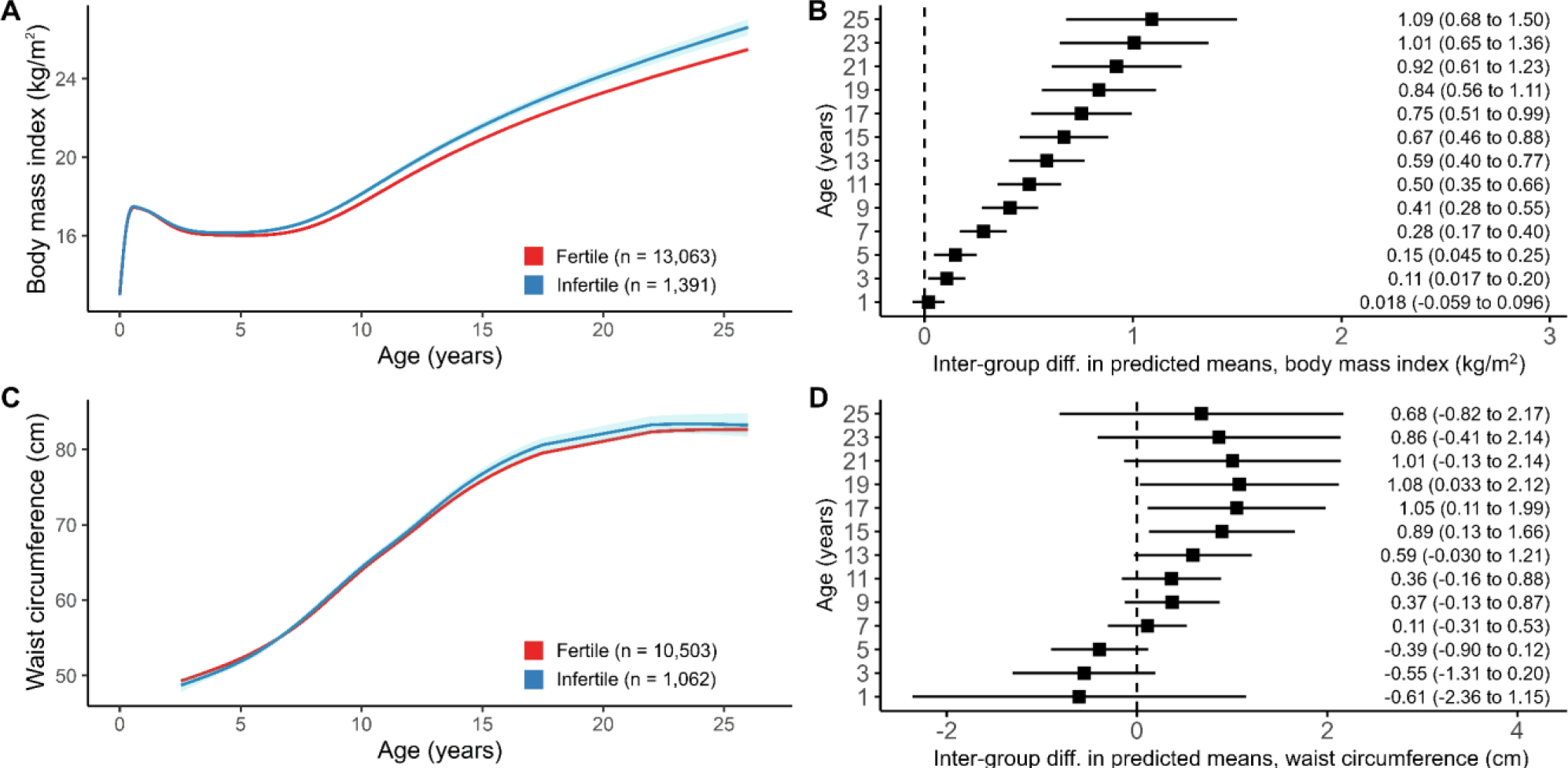
Trajectories of cardiometabolic traits from birth to early adulthood in offspring of couples with (blue) and without infertility (red) and differences in mean predicted values for body mass index (A-B) and waist circumference (C-D).

### Blood pressure

Offspring of couples with infertility had slightly higher mean SBP and DBP values (**Figures 3A** and **3C**). Inter-group differences for SBP peaked around late adolescence (age 17: +0.93 mmHg, 95% CI 0.04 to 1.81, Figure 3B) and those of DBP in early adulthood (age 25: +1.21 mmHg, 95% CI −0.003 to 2.43, Figure 3D).

**Figure 3.**
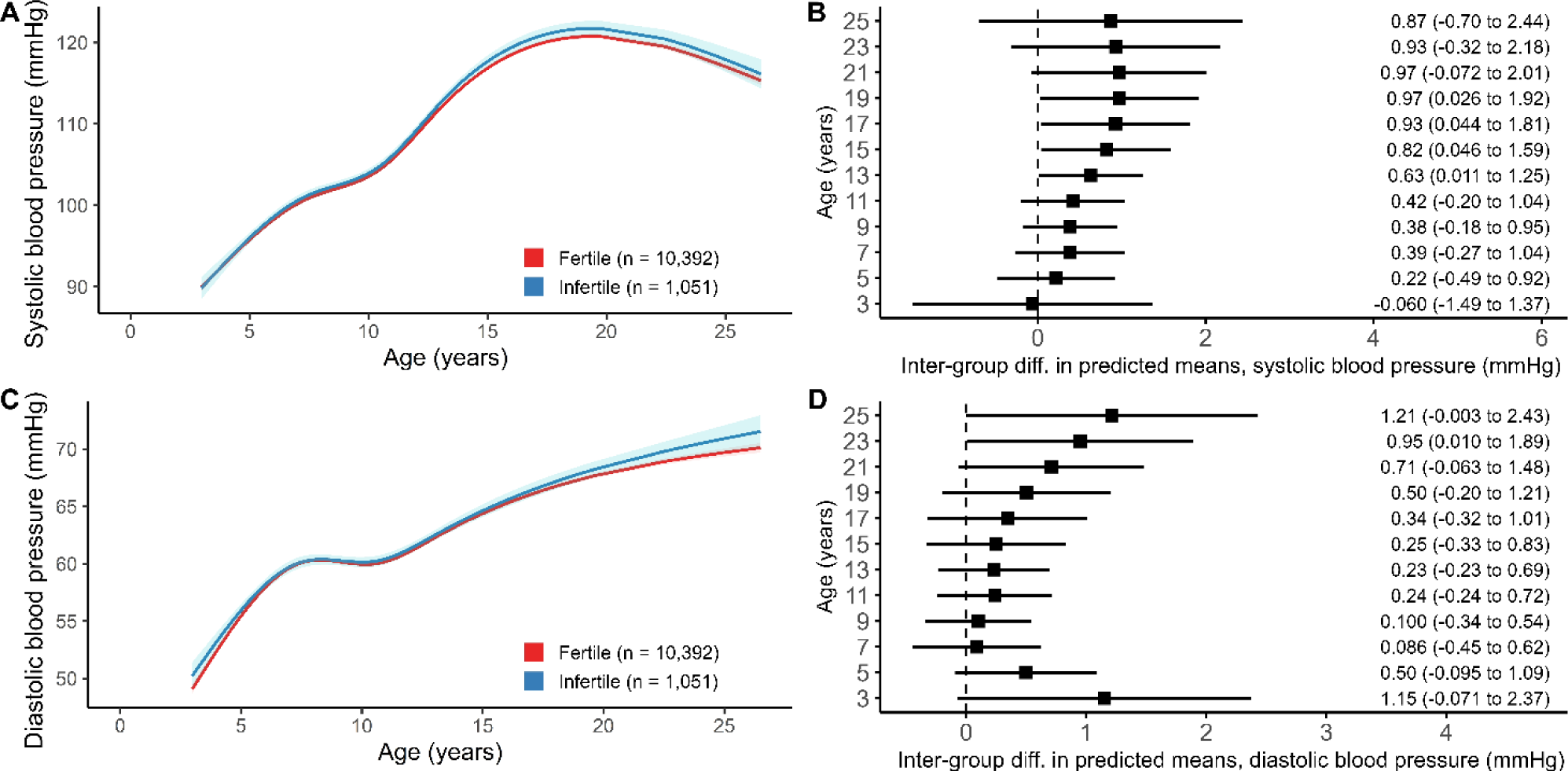
Trajectories of cardiometabolic traits from birth to early adulthood in offspring of couples with (blue) and without infertility (red) and differences in mean predicted values for systolic blood pressure (A-B) and diastolic blood pressure (C-D).

### Lipid profile and glucose

No clear associations between parental infertility and lipid profile and glucose in the offspring were seen. Offspring of couples with infertility had lower average LDL-C during childhood (age 5: −5.04%, 95% CI −8.04 to −2.04), these differences disappeared by age 7, and tended to increase over time from adolescence, which resulted in suggestively higher values in early adulthood (age 25: +4.07%, 95% −0.79 to 8.93, **Figures 4A** and **4B**). Offspring of couples with infertility also tended to have lower HDL-C levels, and this difference increased over time (age 25: −2.78%, 95% CI −6.99 to 1.43, **Figure 4C** and **Figure 4D**). Triglyceride differences between offspring of couples with and without infertility were imprecise, with the largest difference seen during adolescence (age 15: +3.09%, 95% CI −1.24 to 7.42, **Figures 4E** and **4F**). Glucose levels in offspring of infertile parents was only higher in childhood (age 5: +1.00%, 95% CI −0.16 to 2.16) but differences attenuated at later ages (**Figures 4G** and **4H**).

**Figure 4.**
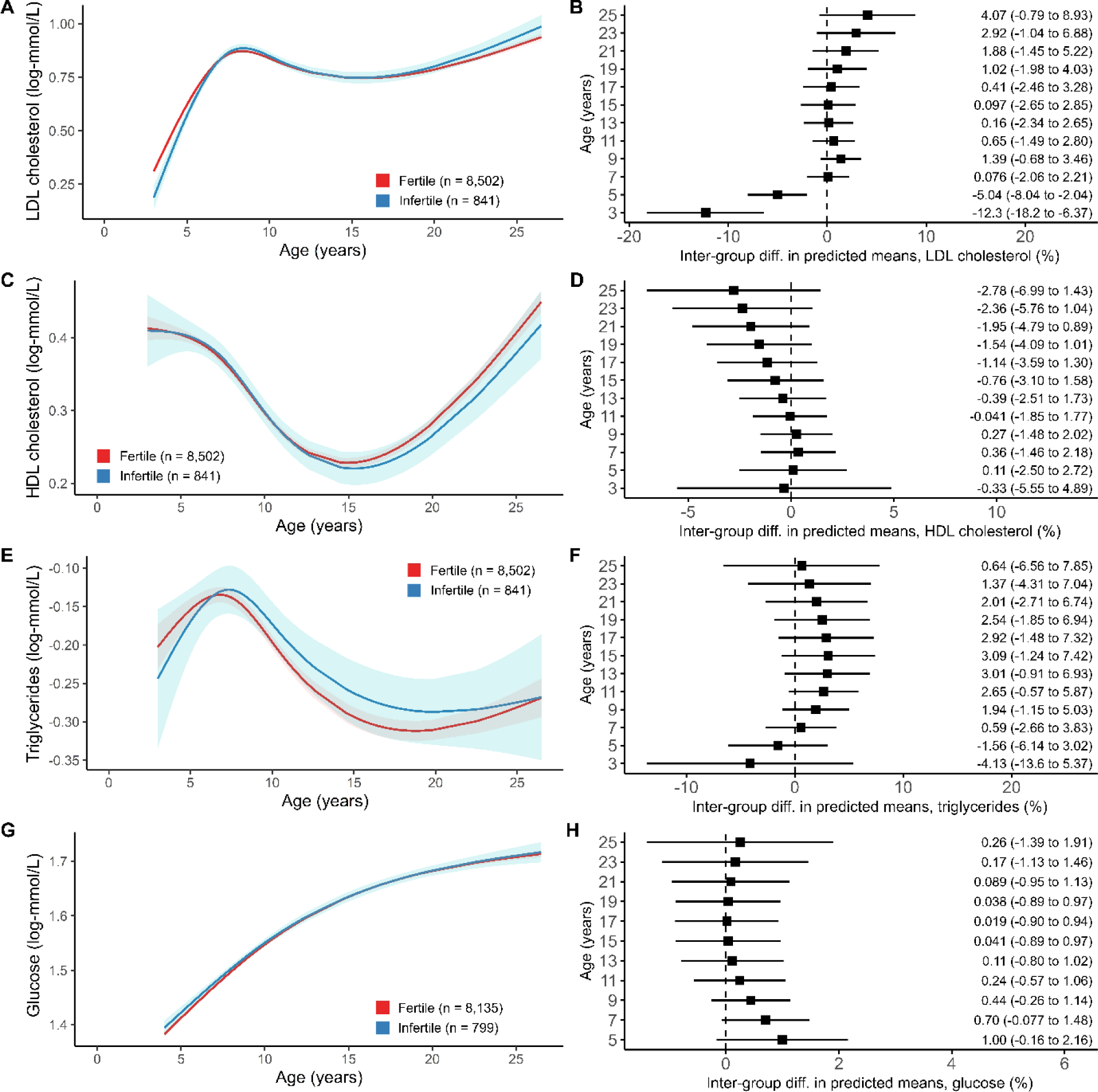
Trajectories of cardiometabolic traits from birth to early adulthood in offspring of couples with (blue) and without infertility (red) and differences in mean predicted values for LDL cholesterol (A-B), HDL cholesterol (C-D), triglycerides (E-F), and glucose (G-H).

### Additional analyses

Further adjustment for the partner’s confounders and both parents’ history of diabetes and hypertension in ALSPAC did not attenuate the inter-group differences in trajectories for any of the cardiometabolic traits (**Supplemental Figure 1**).

We observed no substantial differences in the cohort-specific trajectories for adiposity measures (**Supplemental Figure 2**). We also observed no substantial differences in the cohort-specific trajectories for SBP. However, the DBP trajectories in G21 seemed different to what was observed in the other two cohorts. Nevertheless, inter-group differences in DBP in early adulthood in the main analyses were consistent with those observed for ALSPAC data only (**Supplemental Figure 3**). We further observed no substantial differences in the cohort-specific trajectories for LDL-C and glucose (**Supplemental Figure 4A** and **4D**), but the shape of HDL-C and triglyceride trajectories seemed not comparable across cohorts (**Supplemental Figure 4B** and **4C**).

Cardiometabolic trajectories in the age range for which there is information in the three cohorts was comparable for anthropometric measures, blood pressure, and LDL-C, but not for HDL-C, triglycerides, and glucose (**Supplemental Figure 5**). Including ART in the definition of infertility slightly attenuated the differences in BMI between offspring of couples with infertility and those without but it did not notably change the rest of the results (**Supplemental Figure 6**).

## DISCUSSION

We found that offspring of couples with infertility had higher BMI from childhood to adulthood (differences widened with age and reached their maximum at 25 years, with a difference of 1.1 kg/m^2^), higher WC during adolescence (these differences attenuated in later ages), slightly higher mean SBP and DBP from childhood to adulthood, and tended to have higher LDL-C and lower HDL-C in adulthood (these trends emerged in adolescence and broadened over time). The observed differences were modest but robust to adjustment including for parental risk factors for cardiovascular disease and infertility.

There is limited evidence available on differences in cardiometabolic health according to parental infertility. In a previous study of 4,151 Danish singletons with 1-28 measures of weight between ages 0-20, offspring of couples with infertility showed a greater linear increase in weight per year until age 10 (16). Two other studies (one with 1,773 Danish children with 1 BMI measurement at age 5, one with 79,740 Norwegian children with 12 BMI measurements between ages 0-7) reported no association between parental infertility and offspring BMI (17, 18), which is consisent with our findings of no differences up to age 7. We did not find more studies on the topic that did not involve participants from ALSPAC, G21 or ABCD (the only other study available is based on ABCD data (28)).

Previous bibliography has focused on differences in cardiometabolic health according to conception by ART. The work presented here builds on two previous papers of ours in which we explored the association of ART conception with cardiometabolic traits, cross-sectionally at different ages from infancy to 26 years in all the A.R.T.-HEALTH partnership cohorts, and with cardiometabolic trajectories in a subgroup of the three cohorts included in this study plus a fourth cohort (GUSTO) (14, 15). In the cross-sectional analyses, ART-conceived offspring had slightly lower mean BMI and WC up to age 9 compared to naturally conceived offspring, shifted toward higher adiposity values from age 17 (15), and had higher LDL-C and HDL-C levels and no differences in blood pressure, triglycerides, and glucose (14). In the trajectory analyses, ART-conceived offspring tended to have higher SBP and lower HDL-C over time and had higher glucose during the preschool years (14). Other studies have reported that ART-conceived offspring have lower BMI values under age 5 (29) but a higher risk of obesity during later childhood and adolescence (30) and greater linear increments in weight until age 20 (16). They also have lower blood pressure at age 6 (29) and no increased risk of diabetes during childhood and adolescence (30). Overall, studies on ART-conceived offspring agree with our findings, showing a trend towards higher values of adiposity in adulthood, higher blood pressure, a trend towards lower HDL-C with age, and lack of associations with glucose and triglycerides. According to our results, underlying parental infertility might explain at least part of the differences in cardiometabolic health associated with ART use as differences in cardiometabolic health remained when ART-conceived offspring were excluded from analyses.

There are several potential explanations for the association between parental infertility and poorer cardiometabolic health in the offspring. Individuals with infertility are prone to have impaired cardiometabolic risk factors (7, 8), which may be inherited by their offspring (9). Infertility could be due to underlying conditions in women such as polycystic ovary syndrome, which is also linked to poorer cardiometabolic health in their offspring (31–33). In parallel, a shared detrimental environment (e.g., unhealthy diet, lack of physical activity) could explain both fertility problems in the parents and the differences in cardiometabolic health in the offspring (34, 35).

This work has some limitations. Our exposure (parental infertility) is a couple-dependent measure, and we could not determine whether the fertility problems originated in the mothers, their male partners, or both. Regarding confounding, we a priori acknowledged known/plausible causes of parental infertility and offspring cardiometabolic traits and several sources of residual confounding. Some confounders were assessed during pregnancy (after the exposure), and we had to assume that there was a strong correlation between pre-pregnancy and during-pregnancy measures. Results in ALSPAC adjusting for confounders available in all three cohorts were not different from those after adjusting for the common covariates plus paternal education, smoking and BMI and paternal and maternal hypertension and diabetes. Residual confounding by paternal dyslipidemia may explain some of the associations, particularly those with offspring lipids, as this variable was unavailable. The harmonisation of confounders to the lowest common denominator may have also resulted in additional residual confouding. Another limitation of our study involves the differential age ranges in our cardiometabolic trait data across the three cohorts. For G21 and ABCD, data are limited to ages 3-12, whereas ALSPAC extends up to age 26. Consequently, our findings regarding cardiometabolic trajectories from age 13 to 26 are solely driven by ALSPAC. We assumed that the cardiometabolic trajectories for the three cohorts, given their shared European origin, would align. However, this assumption may not be entirely precise, as there were inconsistencies in the trajectories within each cohort. These discrepancies may be attributed to the limited number of data points for two of the cohorts, which may have affected the construction of polynomial equations and complicated a qualitative comparison between cohorts. Thus, our conclusions beyond the age of 13 may be subject to potential biases, they should be carefully interpretated and validated in further studies incorporating additional time points in adolescence, early adulthood, or later in life. Our results may have also been affected by selection bias, as there were some differences between included and non-included participants (included participants were older, less prone to be smokers, more educated, more likely to be of European origin, and less likely to have more than one offspring). Finally, our study population, primarily of European origin, may limit the generalizability of our findings to populations with similar genetic backgrounds.

## CONCLUSIONS

Offspring of couples with infertility, compared to those of fertile couples, had increasing BMI differences over time, higher WC in adolescence (which was later attenuated), slightly higher blood pressure from childhood to adulthood, and tended to have higher LDL-C levels and lower HDL-C with age (starting in their adolescence). To the best of our knowledge, this is the largest study assessing the differences in cardiometabolic trajectories between birth and early adulthood according to parental infertility. These findings require validation in larger cohorts, measures in older ages, and a better understanding of their mechanisms but suggest that parental infertility may be an indicator of a poorer cardiometabolic profile in offspring.

## Data Availability

Consent given by the participants does not allow individual level participant data to be presented in repositories or journals. Data used in this study is available to bone fide researchers upon request to each cohort (ALSPAC: application to the ALSPAC Executive Committee through https://proposals.epi.bristol.ac.uk/; G21: application to Dr. Henrique Barros; ABCD: application according to the guidelines in the ABCD website [https://www.amc.nl/web/abcd-studie-2/abcd-studie/achtergrondinformatie-abcd-studie.htm], directed to). Please contact Prof. Deborah A. Lawlor and Dr. Ahmed Elhakeem if you have relevant data and would like to join the A.R.T-HEALTH partnership (https://arthealth.bristol.ac.uk/) and contribute to future collaborations.

## ACKNOWLEDGEMENTS

We are extremely grateful to: all the families who took part in the ALSPAC study, the midwives for their help in recruiting them, and the whole ALSPAC team, which includes interviewers, computer and laboratory technicians, clerical workers, research scientists, volunteers, managers, receptionists and nurses; all women who participated in G21 and the whole G21 team; and the hospitals, obstetric clinics, and general practitioners who implemented the ABCD study and all women who participated in ABCD for their cooperation.

## SUPPLEMENTAL MATERIALS

## SUPPLEMENTAL METHODS

### Detailed description of study numbers in ALSPAC

Pregnant women resident in Avon, United Kingdom, with expected dates of delivery between April 1, 1991 and December 31, 1992 were invited to participate in the study. 20,248 pregnancies were identified as eligible and the initial number of pregnancies enrolled was 14,541. Of the initial pregnancies, there was a total of 14,676 fetuses, resulting in 14,062 live births and 13,988 children who were alive at 1 year of age. When the oldest children were approximately 7 years of age, an attempt was made to bolster the initial sample with eligible cases who had failed to join the study originally. As a result, when considering variables collected from the age of seven onwards (and potentially abstracted from obstetric notes), there are data available for more than the 14,541 pregnancies mentioned above. The total sample size for analyses using any data collected after the age of seven is 15,447 pregnancies, resulting in 15,658 fetuses. Of these, 14,901 children were alive at age 1.

Of the original 14,541 initial pregnancies, 338 were from a woman who had already enrolled with a previous pregnancy, meaning 14,203 unique mothers were initially enrolled in the study. As a result of the additional phases of recruitment, a further 630 women who did not enroll originally have provided data since their child was 7 years of age. This provides a total of 14,833 unique women (G0 mothers) enrolled in ALSPAC as of September 2021.

G0 partners were invited to complete questionnaires by the mothers at the start of the study and they were not formally enrolled at that time. 12,113 G0 partners have been in contact with the study by providing data and/or formally enrolling when this started in 2010. 3,807 G0 partners are currently enrolled.

---

**Supplemental Figure 1.**
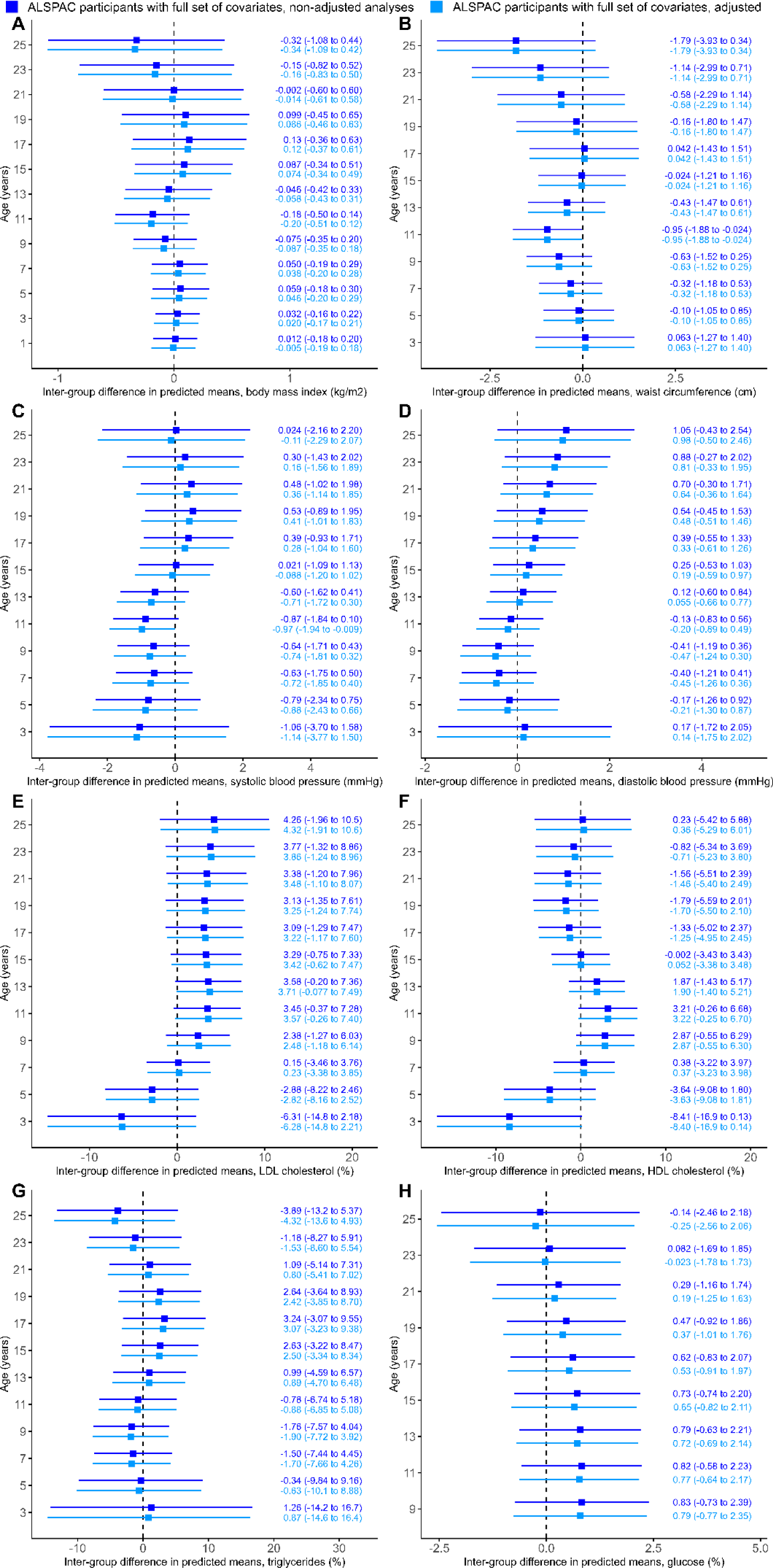
Inter-group differences in ALSPAC participants in mean predicted values for cardiometabolic traits. Dark blue: analyses adjusted for sex of the offspring, cohort, maternal age, maternal BMI, maternal smoking, maternal education, parity, and ethnicity. Light blue: analyses adjusted for the previous confounders + partner’s age, BMI, smoking, and educational level, and both parent’s history of diabetes and hypertension. A: body mass index. B: waist circumference. C: systolic blood pressure. D: diastolic blood pressure. E: LDL cholesterol. F: HDL cholesterol. G: triglycerides. H: glucose.

**Supplemental Figure 2.**
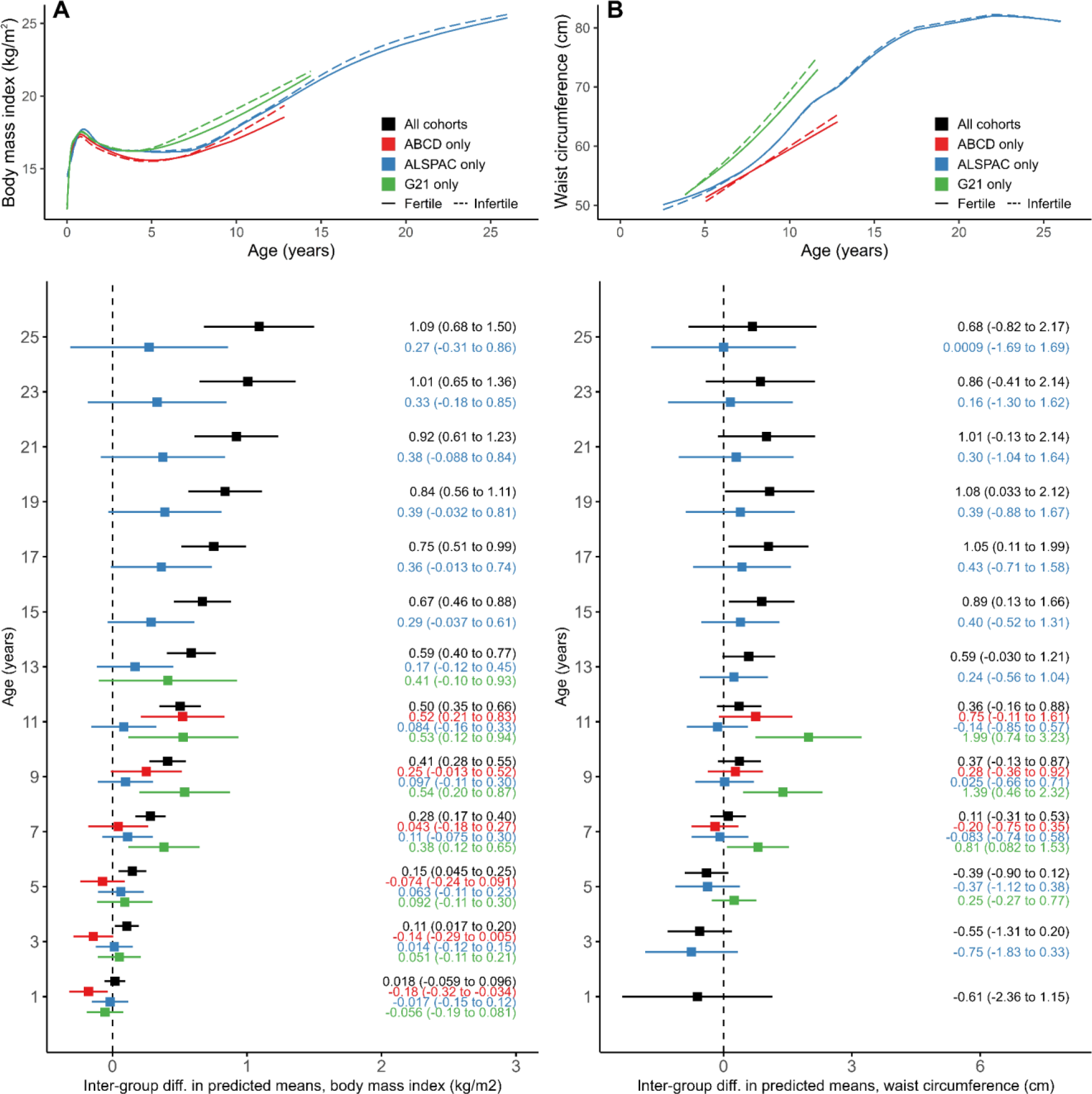
Cohort-specific trajectories (ABCD: red; ALSPAC: blue; G21: green) from childhood to early adulthood in offspring of couples with (continuous line) and without infertility (dashed line), and inter-group differences in mean predicted values for body mass index (A) and waist circumference (B).

**Supplemental Figure 3.**
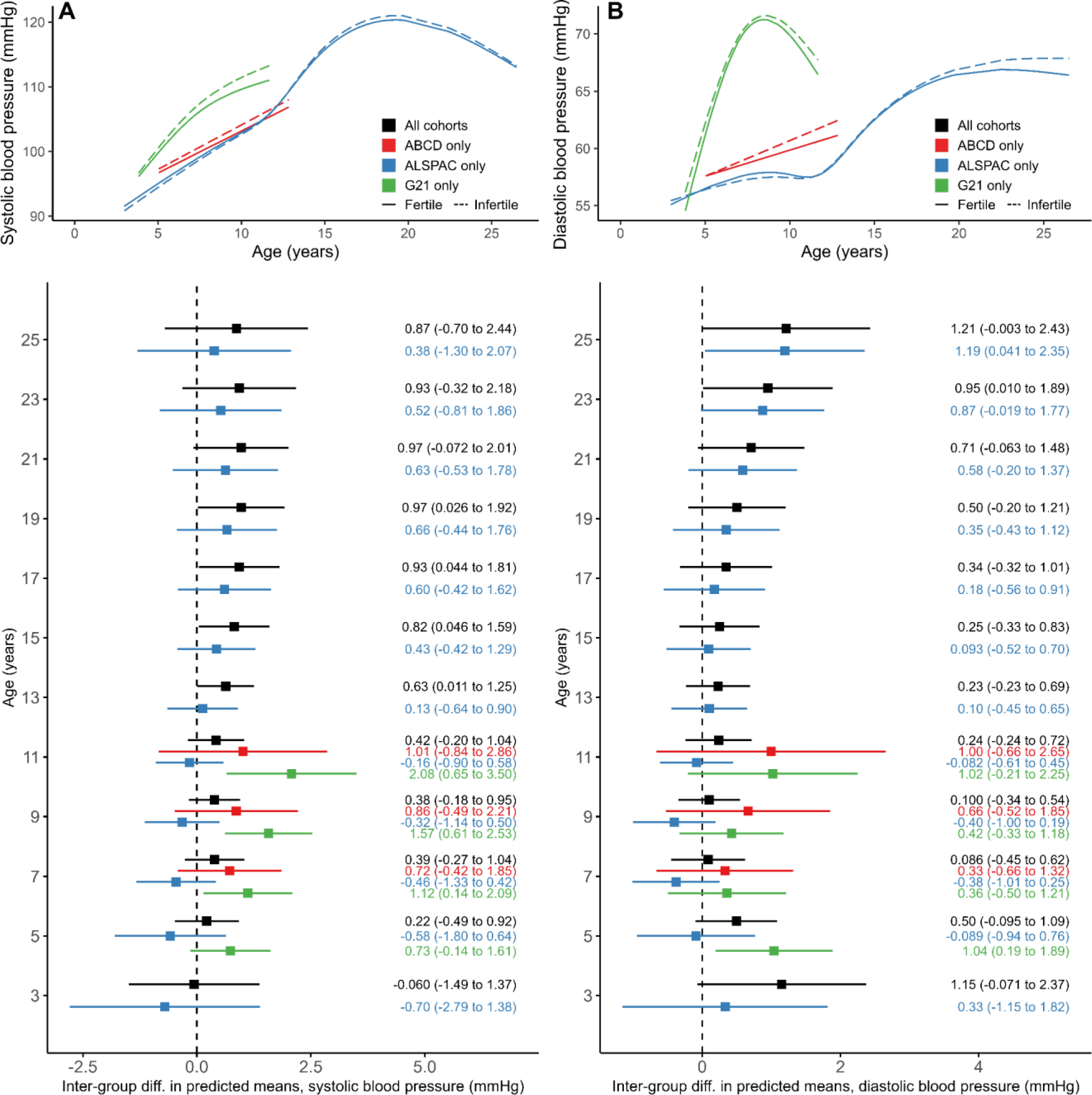
Cohort-specific trajectories (ABCD: red; ALSPAC: blue; G21: green) from childhood to early adulthood in offspring of couples with (continuous line) and without infertility (dashed line), and inter-group differences in mean predicted values for systolic blood pressure (A) and diastolic blood pressure (B).

**Supplemental Figure 4.**
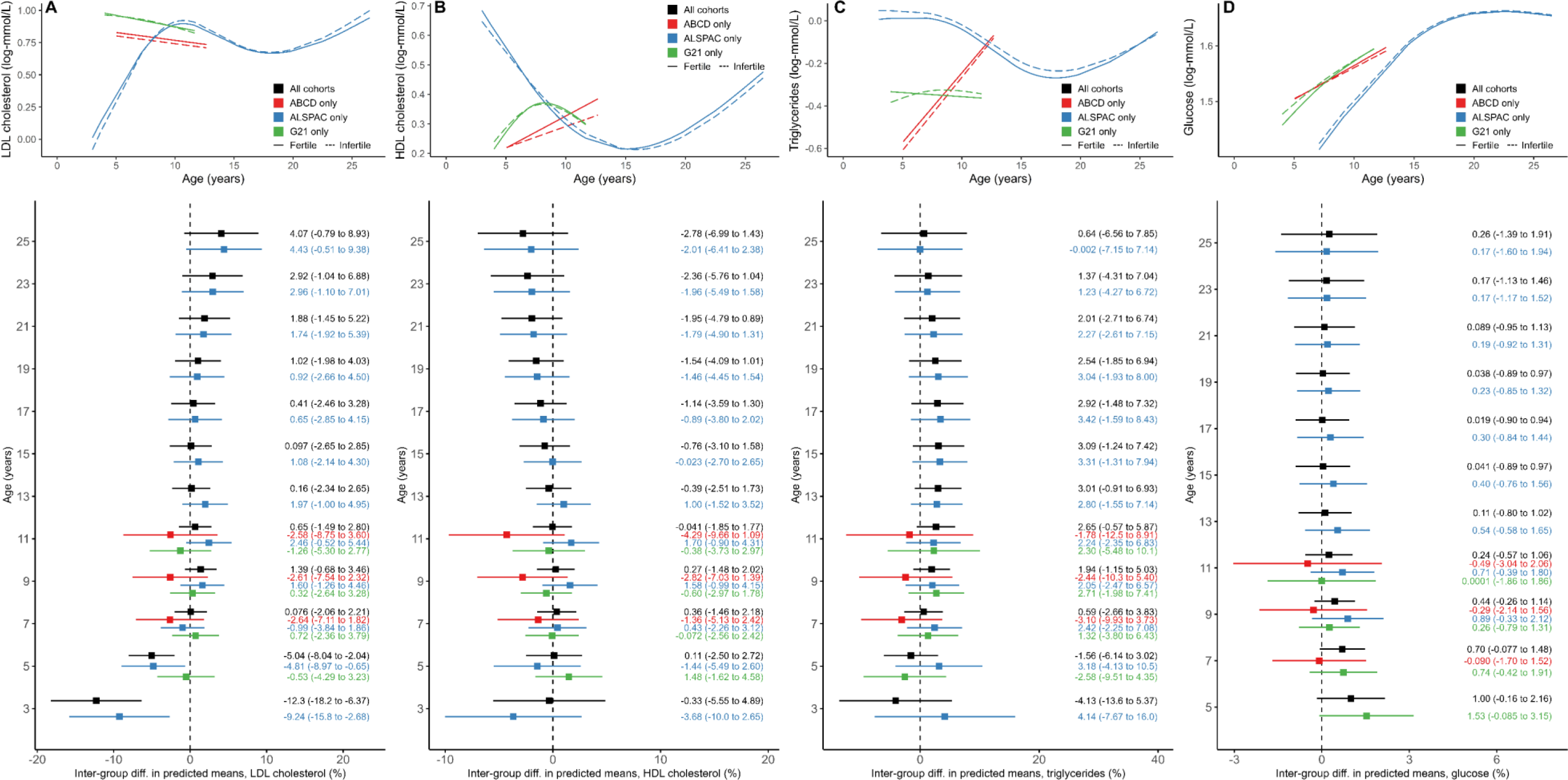
Cohort-specific trajectories (ABCD: red; ALSPAC: blue; G21: green) from childhood to early adulthood in offspring of couples with (continuous line) and without infertility (dashed line), and inter-group differences in mean predicted values for LDL cholesterol (A), HDL cholesterol (B), triglycerides (C), and glucose (D).

**Supplemental Figure 5.**
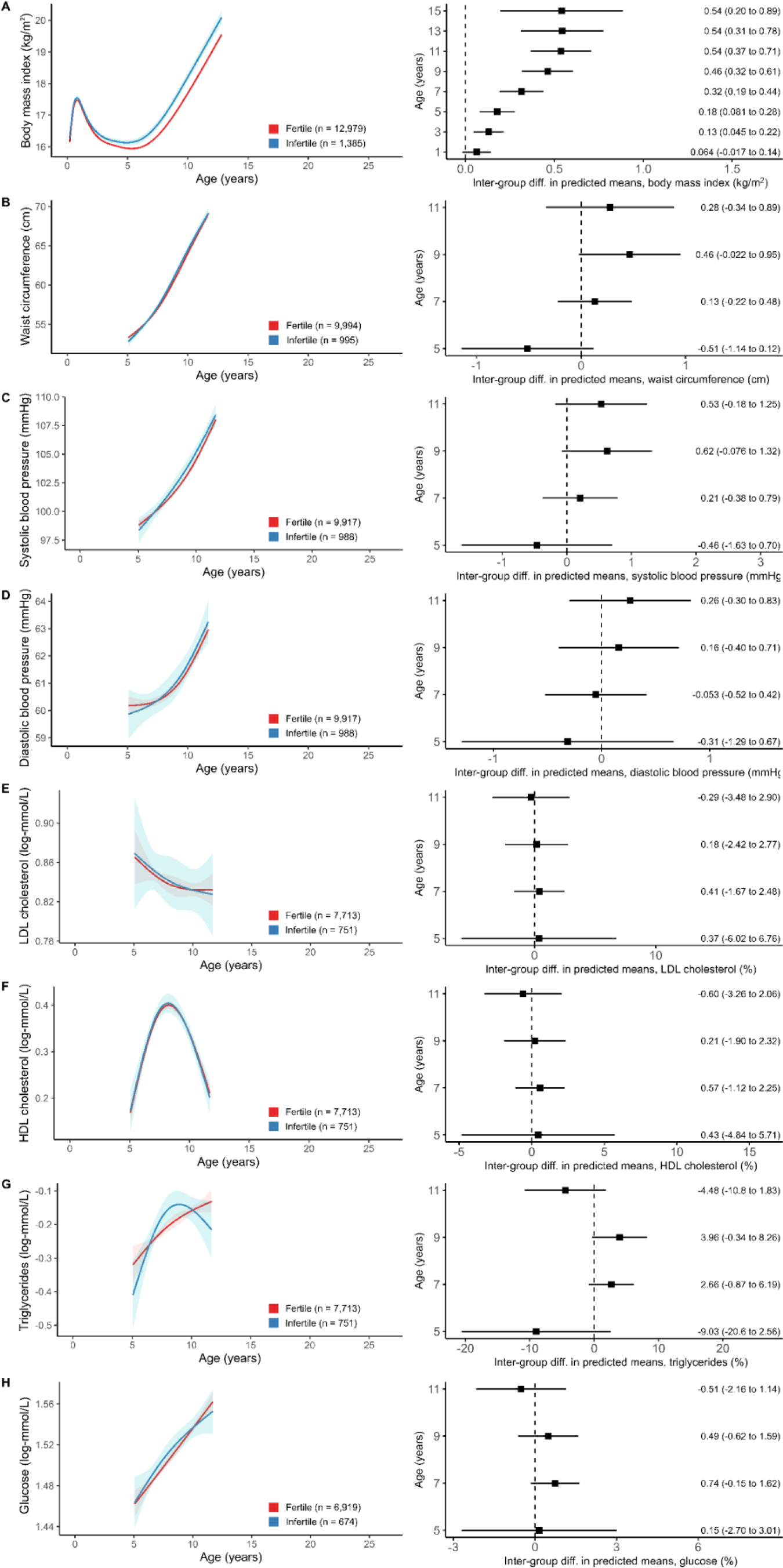
Inter-group differences in mean predicted values for cardiometabolic traits in the age ranges for which there is information in the three cohorts. A: body mass index. B: waist circumference. C: systolic blood pressure. D: diastolic blood pressure. E: LDL cholesterol. F: HDL cholesterol. G: triglycerides. H: glucose.

**Supplemental Figure 6.**
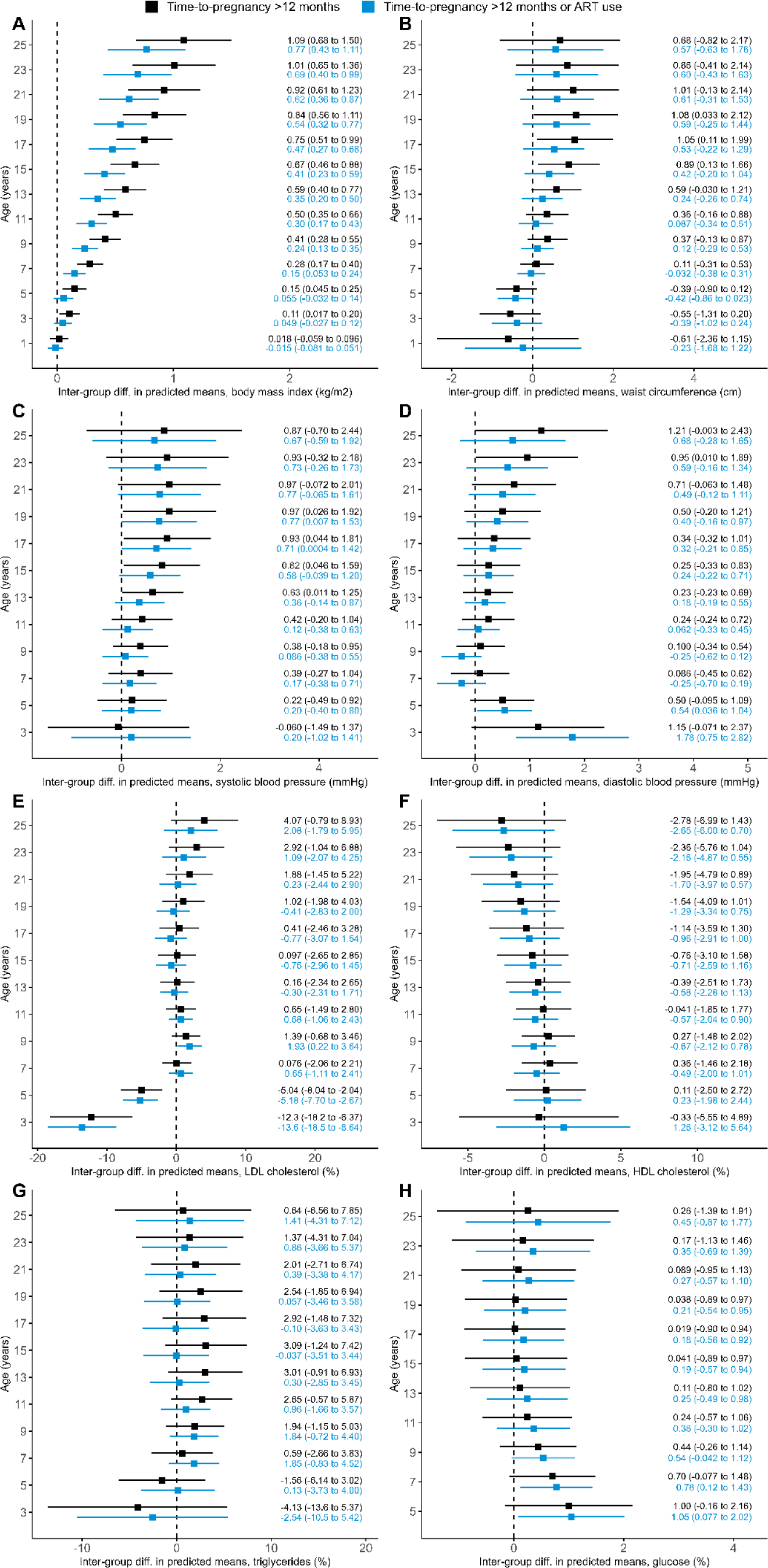
Inter-group differences in mean predicted values for cardiometabolic traits using the standard definition of infertility (time-to-pregnancy > 12 months, black) and the expanded one (time-to-pregnancy > 12 months or ART use, blue). A: body mass index. B: waist circumference. C: systolic blood pressure. D: diastolic blood pressure. E: LDL cholesterol. F: HDL cholesterol. G: triglycerides. H: glucose.

**Supplemental Table 1.** Number of participants with information on each cardiometabolic parameter available in study visits (in increasing order of the average age of the participants).

| Visit | Age, years [mean (SD)] | BMI | WC | Blood pressure | Lipids | Glucose |
| --- | --- | --- | --- | --- | --- | --- |
| **ALSPAC** |  |  |  |  |  |  |
| #1 | 0.1 (0.0) | 6143 | - | - | - | - |
| #2 | 0.3 (0.0) | 611 | - | - | - | - |
| #3 | 0.7 (0.0) | 807 | - | - | - | - |
| #4 | 0.8 (0.1) | 5805 | - | - | - | - |
| #5 | 1.5 (0.0) | 721 | - | - | - | - |
| #6 | 1.7 (0.2) | 5349 | - | - | - | - |
| #7 | 2.1 (0.0) | 656 | - | - | - | - |
| #8 | 2.6 (0.0) | 687 | 690 | - | - | - |
| #9 | 3.1 (0.0) | 663 | 661 | 638 | 216 | - |
| #10 | 3.6 (0.0) | 661 | 657 | - | 383 | - |
| #11 | 3.7 (0.2) | 4954 | - | - | - | - |
| #12 | 4.1 (0.0) | 656 | 654 | 617 | - | - |
| #13 | 5.2 (0.1) | 624 | 619 | 593 | - | - |
| #14 | 7.5 (0.2) | 4683 | 4685 | 4603 | 3087 | 3122 |
| #15 | 8.6 (0.2) | 3944 | - | - | - | - |
| #16 | 9.9 (0.3) | 4370 | 4408 | 4357 | 2906 | - |
| #17 | 10.7 (0.2) | 4311 | 4337 | 4159 | - | - |
| #18 | 11.8 (0.2) | 4123 | 4122 | 4075 | - | - |
| #19 | 12.8 (0.2) | 3892 | 3905 | 3870 | - | - |
| #20 | 13.9 (0.2) | 3618 | 3613 | 3157 | - | - |
| #21 | 15.5 (0.3) | 3172 | 2643 | 3110 | 2038 | 2038 |
| #22 | 17.8 (0.4) | 2933 | - | 2721 | 1910 | 1910 |
| #23 | 24.0 (0.8) | 2330 | 2324 | 2341 | 1913 | 1914 |
| **G21** |  |  |  |  |  |  |
| #1 | 0.1 (0.6) | 3864 | - | - | - | - |
| #2 | 0.2 (0.8) | 3847 | - | - | - | - |
| #3 | 0.3 (0.9) | 3825 | - | - | - | - |
| #4 | 0.5 (1.0) | 3805 | - | - | - | - |
| #5 | 0.6 (1.1) | 3772 | - | - | - | - |
| #6 | 0.8 (1.1) | 3737 | - | - | - | - |
| #7 | 1.0 (1.2) | 3699 | - | - | - | - |
| #8 | 1.2 (1.3) | 3652 | - | - | - | - |
| #9 | 1.4 (1.4) | 3596 | - | - | - | - |
| #10 | 1.8 (1.6) | 3531 | - | - | - | - |
| #11 | 2.2 (1.8) | 3449 | - | - | - | - |
| #12 | 2.6 (2.1) | 3319 | - | - | - | - |
| #13 | 3.1 (2.3) | 3170 | - | - | - | - |
| #14 | 3.6 (2.5) | 2986 | - | - | - | - |
| #15 | 4.1 (2.7) | 2788 | - | - | - | - |
| #16 | 4.3 (0.4) | - | 3394 | 2679 | 900 | 900 |
| #17 | 4.6 (2.8) | 2539 | - | - | - | - |
| #18 | 5.1 (3.0) | 2305 | - | - | - | - |
| #19 | 5.6 (3.0) | 2055 | - | - | - | - |
| #20 | 5.9 (3.0) | 1803 | - | - | - | - |
| #21 | 6.4 (3.0) | 1587 | - | - | - | - |
| #22 | 6.8 (3.0) | 1376 | - | - | - | - |
| #23 | 7.1 (0.2) | - | 3371 | 3364 | 2607 | 2617 |
| #24 | 7.2 (3.0) | 1164 | - | - | - | - |
| #25 | 7.6 (3.0) | 974 | - | - | - | - |
| #26 | 7.8 (2.9) | 796 | - | - | - | - |
| #27 | 8.2 (2.9) | 652 | - | - | - | - |
| #28 | 8.4 (2.9) | 503 | - | - | - | - |
| #29 | 8.7 (2.9) | 392 | - | - | - | - |
| #30 | 8.9 (2.8) | 297 | - | - | - | - |
| #31 | 9.0 (2.5) | 14 | - | - | - | - |
| #32 | 9.1 (2.7) | 217 | - | - | - | - |
| #33 | 9.2 (2.8) | 50 | - | - | - | - |
| #34 | 9.3 (2.7) | 76 | - | - | - | - |
| #35 | 9.3 (2.8) | 162 | - | - | - | - |
| #36 | 9.4 (2.8) | 117 | - | - | - | - |
| #37 | 9.6 (2.3) | 12 | - | - | - | - |
| #38 | 9.6 (2.8) | 36 | - | - | - | - |
| #39 | 9.9 (1.5) | 8 | - | - | - | - |
| #40 | 10.0 (2.8) | 28 | - | - | - | - |
| #41 | 10.1 (0.3) | - | 3143 | 3146 | 2217 | 2234 |
| #42 | 11.0 (1.6) | 8 | - | - | - | - |
| #43 | 11.6 (0.9) | 6 | - | - | - | - |
| #44 | 12.1 (0.6) | 4 | - | - | - | - |
| #45 | 12.3 (0.6) | 2 | - | - | - | - |
| **ABCD** |  |  |  |  |  |  |
| #1 | 0.4 (0.6) | 2704 | - | - | - | - |
| #2 | 0.4 (0.7) | 1930 | - | - | - | - |
| #3 | 0.5 (0.5) | 2872 | - | - | - | - |
| #4 | 0.6 (0.4) | 2880 | - | - | - | - |
| #5 | 0.8 (0.5) | 2867 | - | - | - | - |
| #6 | 0.8 (1.1) | 595 | - | - | - | - |
| #7 | 1.1 (0.6) | 2779 | - | - | - | - |
| #8 | 1.3 (1.0) | 257 | - | - | - | - |
| #9 | 1.5 (0.9) | 2684 | - | - | - | - |
| #10 | 2.0 (1.1) | 2621 | - | - | - | - |
| #11 | 2.7 (1.3) | 2476 | - | - | - | - |
| #12 | 3.3 (1.4) | 2143 | - | - | - | - |
| #13 | 3.8 (1.4) | 1623 | - | - | - | - |
| #14 | 4.1 (1.4) | 1076 | - | - | - | - |
| #15 | 4.1 (2.2) | 69 | - | - | - | - |
| #16 | 4.3 (1.0) | 7 | - | - | - | - |
| #17 | 4.4 (1.5) | 78 | - | - | - | - |
| #18 | 4.5 (1.4) | 636 | - | - | - | - |
| #19 | 4.5 (1.4) | 50 | - | - | - | - |
| #20 | 4.5 (1.4) | 336 | - | - | - | - |
| #21 | 4.7 (1.4) | 162 | - | - | - | - |
| #22 | 4.8 (1.3) | 19 | - | - | - | - |
| #23 | 5.0 (1.0) | 4 | - | - | - | - |
| #24 | 5.8 (0.5) | 2089 | 2087 | 2014 | 1373 | 1373 |
| #25 | 10.6 (0.4) | 1870 | - | - | - | - |
| #26 | 11.8 (0.4) | 711 | 712 | 654 | 577 | 579 |

**Supplemental Table 2.**
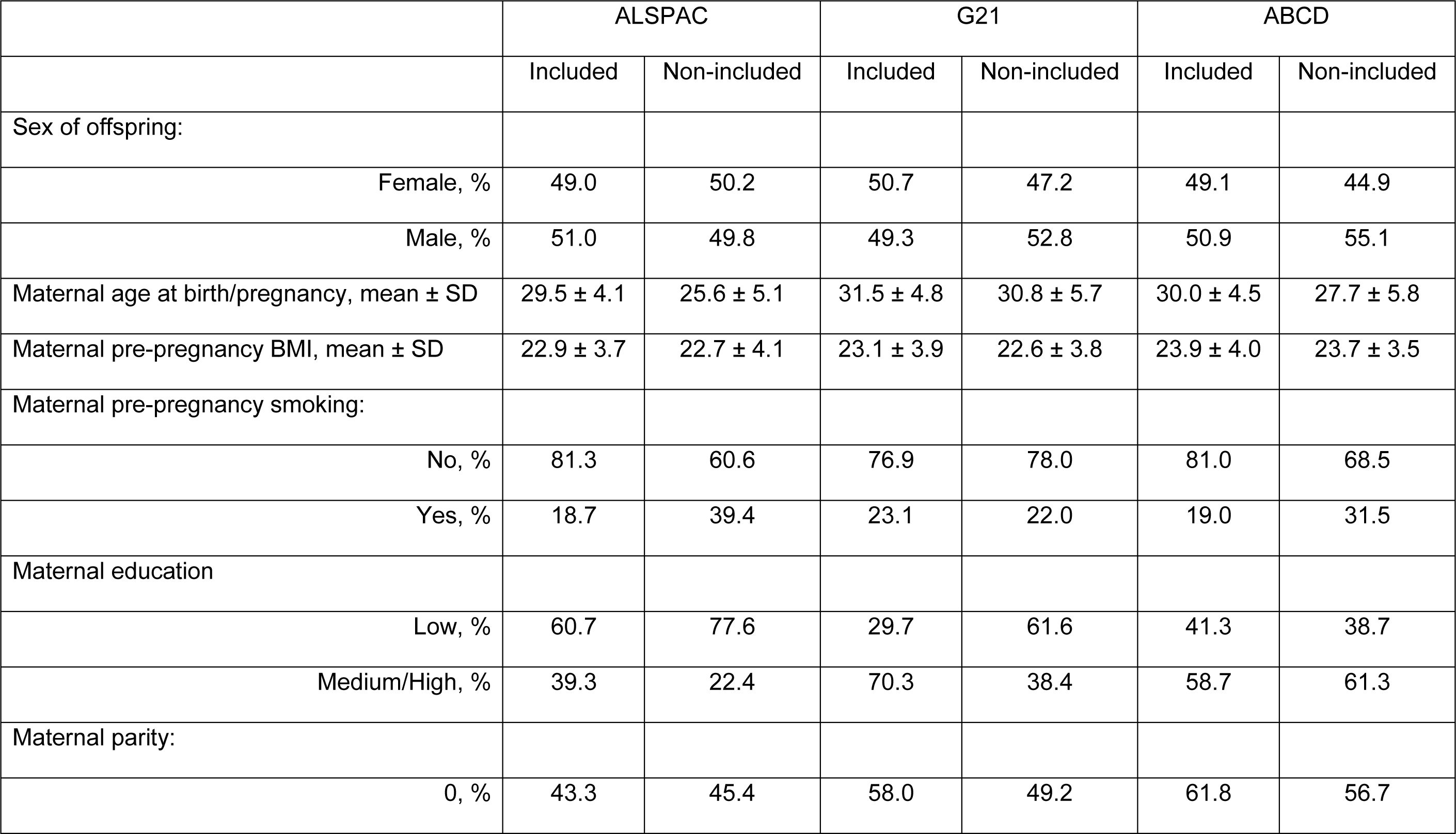
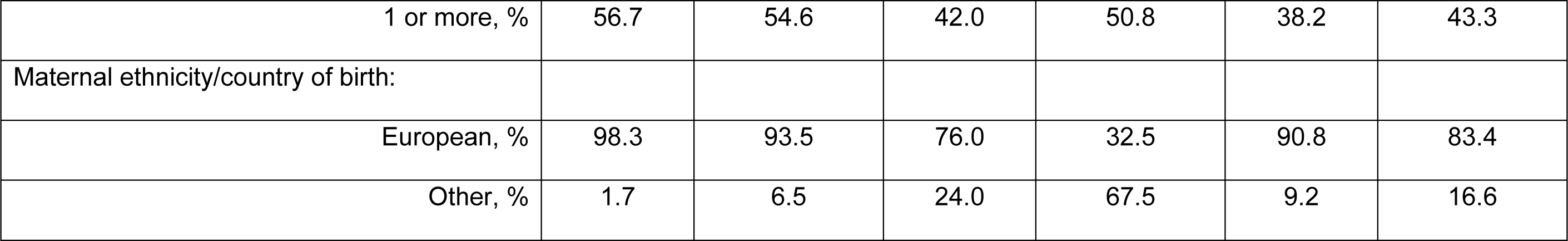
Differences in included versus non-included participants.

|  | ALSPAC |  | G21 |  | ABCD |  |
| --- | --- | --- | --- | --- | --- | --- |
|  | Included | Non-included | Included | Non-included | Included | Non-included |
| Sex of offspring: |  |  |  |  |  |  |
| Female, % | 49.0 | 50.2 | 50.7 | 47.2 | 49.1 | 44.9 |
| Male, % | 51.0 | 49.8 | 49.3 | 52.8 | 50.9 | 55.1 |
| Maternal age at birth/pregnancy, mean ± SD | 29.5 ± 4.1 | 25.6 ± 5.1 | 31.5 ± 4.8 | 30.8 ± 5.7 | 30.0 ± 4.5 | 27.7 ± 5.8 |
| Maternal pre-pregnancy BMI, mean ± SD | 22.9 ± 3.7 | 22.7 ± 4.1 | 23.1 ± 3.9 | 22.6 ± 3.8 | 23.9 ± 4.0 | 23.7 ± 3.5 |
| Maternal pre-pregnancy smoking: |  |  |  |  |  |  |
| No, % | 81.3 | 60.6 | 76.9 | 78.0 | 81.0 | 68.5 |
| Yes, % | 18.7 | 39.4 | 23.1 | 22.0 | 19.0 | 31.5 |
| Maternal education |  |  |  |  |  |  |
| Low, % | 60.7 | 77.6 | 29.7 | 61.6 | 41.3 | 38.7 |
| Medium/High, % | 39.3 | 22.4 | 70.3 | 38.4 | 58.7 | 61.3 |
| Maternal parity: |  |  |  |  |  |  |
| 0, % | 43.3 | 45.4 | 58.0 | 49.2 | 61.8 | 56.7 |
| 1 or more, % | 56.7 | 54.6 | 42.0 | 50.8 | 38.2 | 43.3 |
| Maternal ethnicity/country of birth: |  |  |  |  |  |  |
| European, % | 98.3 | 93.5 | 76.0 | 32.5 | 90.8 | 83.4 |
| Other, % | 1.7 | 6.5 | 24.0 | 67.5 | 9.2 | 16.6 |

## REFERENCES

1. Magnus MC, Fraser A, Rich-Edwards JW, Magnus P, Lawlor DA, Håberg SE. Time-to-pregnancy and risk of cardiovascular disease among men and women. Eur J Epidemiol 2021;36:383–91.

2. Skåra KH, Åsvold BO, Hernáez Á, Fraser A, Rich-Edwards JW, Farland LV et al. Risk of cardiovascular disease in women and men with subfertility: the Trøndelag Health Study. Fertility and Sterility 2022.

3. Farland LV, Wang YX, Gaskins AJ, Rich-Edwards JW, Wang S, Magnus MC et al. Infertility and Risk of Cardiovascular Disease: A Prospective Cohort Study. J Am Heart Assoc 2023;12:e027755.

4. Murugappan G, Leonard SA, Farland LV, Lau ES, Shadyab AH, Wild RA et al. Association of infertility with atherosclerotic cardiovascular disease among postmenopausal participants in the Women’s Health Initiative. Fertil Steril 2022;117:1038–46.

5. Gleason JL, Shenassa ED, Thoma ME. Self-reported infertility, metabolic dysfunction, and cardiovascular events: a cross-sectional analysis among U.S. women. Fertil Steril 2019;111:138–46.

6. Chen PC, Chen YJ, Yang CC, Lin TT, Huang CC, Chung CH et al. Male Infertility Increases the Risk of Cardiovascular Diseases: A Nationwide Population-Based Cohort Study in Taiwan. World J Mens Health 2022;40:490–500.

7. Mahalingaiah S, Sun F, Cheng JJ, Chow ET, Lunetta KL, Murabito JM. Cardiovascular risk factors among women with self-reported infertility. Fertil Res Pract 2017;3:7.

8. Choy JT, Eisenberg ML. Male infertility as a window to health. Fertil Steril 2018;110:810–4.

9. Hanson M. The inheritance of cardiovascular disease risk. Acta Paediatr 2019;108:1747–56.

10. Tobias DK, Chavarro JE, Williams MA, Buck Louis GM, Hu FB, Rich-Edwards J et al. History of infertility and risk of gestational diabetes mellitus: a prospective analysis of 40,773 pregnancies. Am J Epidemiol 2013;178:1219–25.

11. Sibai BM. Subfertility/infertility and assisted reproductive conception are independent risk factors for pre-eclampsia. Bjog 2015;122:923.

12. Hoodbhoy Z, Mohammed N, Nathani KR, Sattar S, Chowdhury D, Maskatia S et al. The Impact of Maternal Preeclampsia and Hyperglycemia on the Cardiovascular Health of the Offspring: A Systematic Review and Meta-Analysis. Am J Perinatol 2021.

13. Pathirana MM, Lassi ZS, Roberts CT, Andraweera PH. Cardiovascular risk factors in offspring exposed to gestational diabetes mellitus in utero: systematic review and meta-analysis. J Dev Orig Health Dis 2020;11:599–616.

14. Elhakeem A, Taylor AE, Inskip HM, Huang JY, Mansell T, Rodrigues C et al. Long-term cardiometabolic health in people born after assisted reproductive technology: a multi-cohort analysis. Eur Heart J 2023.

15. Elhakeem A, Taylor AE, Inskip HM, Huang J, Tafflet M, Vinther JL et al. Association of Assisted Reproductive Technology With Offspring Growth and Adiposity From Infancy to Early Adulthood. JAMA Netw Open 2022;5:e2222106.

16. Hinchely Ebdrup N, Hohwü L, Bay B, Obel C, Knudsen UB, Kesmodel US. Long-term growth in offspring of infertile parents: A 20-year follow-up study. Acta Obstet Gynecol Scand 2021;100:1849–57.

17. Bay B, Mortensen EL, Kesmodel US. Is subfertility or fertility treatment associated with long-term growth in the offspring? A cohort study. Fertil Steril 2014;102:1117–23.

18. Magnus MC, Wilcox AJ, Fadum EA, Gjessing HK, Opdahl S, Juliusson PB et al. Growth in children conceived by ART. Hum Reprod 2021;36:1074–82.

19. Boyd A, Golding J, Macleod J, Lawlor DA, Fraser A, Henderson J et al. Cohort Profile: the ‘children of the 90s’--the index offspring of the Avon Longitudinal Study of Parents and Children. Int J Epidemiol 2013;42:111–27.

20. Fraser A, Macdonald-Wallis C, Tilling K, Boyd A, Golding J, Davey Smith G et al. Cohort Profile: the Avon Longitudinal Study of Parents and Children: ALSPAC mothers cohort. Int J Epidemiol 2013;42:97–110.

21. Major-Smith D, Heron J, Fraser A, Lawlor D, Golding J, Northstone K. The Avon Longitudinal Study of Parents and Children (ALSPAC): a 2022 update on the enrolled sample of mothers and the associated baseline data [version 2; peer review: 2 approved]. Wellcome Open Research 2023;7.

22. Northstone K, Ben Shlomo Y, Teyhan A, Hill A, Groom A, Mumme M et al. The Avon Longitudinal Study of Parents and children ALSPAC G0 Partners: A cohort profile [version 2; peer review: 1 approved]. Wellcome Open Research 2023;8.

23. Northstone K, Lewcock M, Groom A, Boyd A, Macleod J, Timpson N et al. The Avon Longitudinal Study of Parents and Children (ALSPAC): an update on the enrolled sample of index children in 2019 [version 1; peer review: 2 approved]. Wellcome Open Research 2019;4.

24. Araújo FA, Lucas R, Simpkin AJ, Heron J, Alegrete N, Tilling K et al. Associations of anthropometry since birth with sagittal posture at age 7 in a prospective birth cohort: the Generation XXI Study. BMJ open 2017;7:e013412.

25. van Eijsden M, Vrijkotte TG, Gemke RJ, van der Wal MF. Cohort profile: the Amsterdam Born Children and their Development (ABCD) study. Int J Epidemiol 2011;40:1176–86.

26. Harris PA, Taylor R, Thielke R, Payne J, Gonzalez N, Conde JG. Research electronic data capture (REDCap)--a metadata-driven methodology and workflow process for providing translational research informatics support. J Biomed Inform 2009;42:377–81.

27. Elhakeem A, Hughes RA, Tilling K, Cousminer DL, Jackowski SA, Cole TJ et al. Using linear and natural cubic splines, SITAR, and latent trajectory models to characterise nonlinear longitudinal growth trajectories in cohort studies. BMC Med Res Methodol 2022;22:68.

28. Pontesilli M, Painter RC, Grooten IJ, van der Post JA, Mol BW, Vrijkotte TG et al. Subfertility and assisted reproduction techniques are associated with poorer cardiometabolic profiles in childhood. Reprod Biomed Online 2015;30:258–67.

29. Huang JY, Cai S, Huang Z, Tint MT, Yuan WL, Aris IM et al. Analyses of child cardiometabolic phenotype following assisted reproductive technologies using a pragmatic trial emulation approach. Nat Commun 2021;12:5613.

30. Norrman E, Petzold M, Gissler M, Spangmose AL, Opdahl S, Henningsen AK et al. Cardiovascular disease, obesity, and type 2 diabetes in children born after assisted reproductive technology: A population-based cohort study. PLoS Med 2021;18:e1003723.

31. Chen X, Koivuaho E, Piltonen TT, Gissler M, Lavebratt C. Association of maternal polycystic ovary syndrome or anovulatory infertility with obesity and diabetes in offspring: a population-based cohort study. Hum Reprod 2021;36:2345–57.

32. Gunning MN, Sir Petermann T, Crisosto N, van Rijn BB, de Wilde MA, Christ JP et al. Cardiometabolic health in offspring of women with PCOS compared to healthy controls: a systematic review and individual participant data meta-analysis. Hum Reprod Update 2020;26:103–17.

33. Wei SQ, Bilodeau-Bertrand M, Auger N. Association of PCOS with offspring morbidity: a longitudinal cohort study. Hum Reprod 2022;37:2135–42.

34. Aoun A, Khoury VE, Malakieh R. Can Nutrition Help in the Treatment of Infertility? Prev Nutr Food Sci 2021;26:109–20.

35. Xie F, You Y, Guan C, Gu Y, Yao F, Xu J. Association between physical activity and infertility: a comprehensive systematic review and meta-analysis. J Transl Med 2022;20:237.

